# Galantamine for routine neuromuscular block reversal and treatment of emergence delirium in children after ENT surgery: a retrospective observational study

**DOI:** 10.64898/2026.11.12.26367732

**Authors:** Andrey A. Neverov, Ekaterina A. Kairysh

## Abstract

**Background:** Emergence delirium is a common complication of general anesthesia in children. Preschool age and ear, nose and throat (ENT) procedures are risk factors [1, 2, 3, 4]. The hypothesis linking delirium to central cholinergic deficiency provides a rationale for studying galantamine, an acetylcholinesterase inhibitor that crosses the blood–brain barrier and is used to reverse neuromuscular blockade. We assessed the association between its routine administration and emergence delirium in children after ENT surgery under sevoflurane anesthesia.

**Objective:** To compare emergence delirium frequency and Pediatric Anesthesia Emergence Delirium (PAED) scores after elective ENT surgery with and without routine reversal of neuromuscular blockade with galantamine, and to describe the course of delirium episodes following therapeutic administration.

**Methods:** This retrospective observational study included 207 children aged 2–17 years who underwent elective ENT surgery, predominantly adenoidectomy, tonsillotomy or tonsillectomy, under sevoflurane-based general anesthesia between October 2025 and January 2026. Data were extracted retrospectively from medical records into a study worksheet. PAED scores were checked against clinical descriptions and corrected where discrepancies were identified. The groups reflected the practices of two anesthesiologists. All children in the prophylactic group (n = 102) received galantamine to reverse neuromuscular blockade. In the group without prophylactic administration (n = 105), routine reversal was not performed; galantamine was administered as needed for impaired emergence or suspected residual neuromuscular blockade. The analysis used assessments performed after extubation in the operating room, before treatment for impaired emergence. The primary outcome was PAED > 10, interpreted alongside the clinical descriptions.

**Results:** *Prophylactic use:* PAED > 10 was recorded in 0 of 102 children receiving routine reversal and 19 of 105 without prophylactic administration (18.1%, p < 0.0001). Median PAED scores were 7 [6–8] and 8 [6–9], respectively (p = 0.0015).

*Therapeutic use:* Intravenous galantamine (2.5–10 mg) was administered in 17 episodes of delirium with motor agitation and impaired interaction. Resolution was documented in all 17 cases. Reduced agitation was accompanied by restoration of meaningful interaction. No sedatives were used to treat delirium. With ondansetron and dexamethasone prophylaxis, emetic events were recorded during the first postoperative hour in 17.6% and 21.9% of children, respectively (p = 0.49). No serious adverse events were documented.

**Conclusion:** No PAED scores > 10 were recorded with routine reversal of neuromuscular blockade using galantamine. Following therapeutic administration, delirium resolution was accompanied by restoration of meaningful interaction without sedatives. These retrospective observational findings support further investigation of prophylactic and therapeutic galantamine use. The observations of restored interaction prompted the hypothesis that galantamine could act as an agent promoting emergence from anesthesia.

## INTRODUCTION

Emergence delirium is an acute disturbance of consciousness and orientation during recovery from general anesthesia, often accompanied by nonpurposeful agitation [5, 6]. Estimates of its frequency in children vary considerably because of differences in diagnostic criteria, assessment timing, and the overlapping use of the terms delirium and agitation [6, 7, 8, 9].

The mechanisms of emergence delirium remain unclear; the roles of pain, preoperative anxiety and rapid emergence from sevoflurane anesthesia have been discussed [7, 8, 9].

Galantamine reversibly inhibits acetylcholinesterase, crosses the blood–brain barrier and is used to reverse neuromuscular blockade [10]. These properties prompted interest in its potential effects on emergence from anesthesia.

The neuromuscular block reversal effects of galantamine were described by M. D. Mashkovsky and R. P. Kruglikova-Lvova in 1951 [11]. A. Derezhian and E. Krusteva (1960) and E. Stoyanov and S. Vulchanova (1963) studied Nivalin for this purpose [12, 13]. A possible central effect is reflected in L. Wislicki’s observations of restored interaction (1967) and E. Chakalova and colleagues’ observations of increased wakefulness (1987) [14, 15].

Central anticholinergic syndrome (CAS) is associated with reduced central cholinergic activity and is considered a possible cause of impaired emergence [16, 17, 18, 19, 20]. In 1977, A. Baraka and S. Harik described reversal of the central effects of scopolamine with galantamine [21]. M. S. Danilov and K. M. Lebedinskii (2015) proposed that CAS may reflect excessive or prolonged suppression of central cholinergic activity during general anesthesia [20].

The cholinergic hypothesis of delirium [22] and observations in CAS provide a rationale for studying galantamine, but its relationship with emergence delirium in children remains insufficiently investigated.

In a study of adults by M. S. Danilov and colleagues (2018), CAS was less frequent with galantamine-mediated reversal of neuromuscular blockade than without reversal. This comparison was not randomized, and the comatose form of the syndrome predominated [10].

In 2022, uncertainty about supplies of sugammadex (Bridion) prompted us to use galantamine to reverse neuromuscular blockade when residual muscle weakness was suspected. A stock of 50 vials of Bridion usually lasted 1–1.5 years; this informed the initial order of 50 ampoules of Nivalin (Sopharma, Bulgaria, 5 mg/mL). We subsequently switched to Galantamine (VIFITECH, Russia, 5 mg/mL) and routine reversal of neuromuscular blockade [23, 24].

During the initial period of galantamine use for neuromuscular block reversal, we noticed that children more often emerged oriented and able to interact. This observation prompted us to test the hypothesis that galantamine could prevent emergence delirium.

The novel contribution of this study is the description of two aspects of galantamine use during emergence from anesthesia in children: the absence of documented delirium following prophylactic administration, and resolution of established episodes following therapeutic administration, accompanied by restoration of meaningful interaction without sedatives.

The study aimed to compare emergence delirium frequency and PAED scores in children after elective ENT surgery with and without routine reversal of neuromuscular blockade with galantamine, and to describe the course of delirium episodes following therapeutic administration.

## MATERIALS AND METHODS

The study was conducted at Krasnoyarsk Interdistrict Children’s Hospital No. 4. Data were obtained from the medical records of patients who underwent elective ENT procedures from 28 October 2025 to 29 January 2026. The study worksheet was a summary table compiled retrospectively by extracting data from these records. The design was retrospective, observational, nonrandomized and unblinded. Inclusion criteria were age 2–17 years, elective ENT surgery, and sevoflurane-based general anesthesia with rocuronium. Exclusion criteria were emergency surgery, American Society of Anesthesiologists (ASA) physical status III or higher, a history of neurological or psychiatric disease, and perioperative dexmedetomidine use. All 207 patients recorded in the worksheet were included in the analysis. No a priori sample size calculation was performed; sample size was determined by the number of available records during the observation period. Based on our clinical estimates, anesthesia lasted approximately 30–40 minutes; duration was not recorded as a separate variable in the analytical dataset.

Adenoidectomy, tonsillotomy and tonsillectomy predominated. The sample also included children undergoing septoplasty, ear surgery and other elective ENT procedures.

The basic anesthetic regimen was the same in both groups. Atropine was not used. No premedication was given for adenoidectomy; midazolam was used in selected adolescents aged 14–17 years before longer procedures. Midazolam premedication was not a reason for exclusion and was not recorded as a separate variable.

### Induction

inhalational induction with sevoflurane by face mask, fentanyl 1 µg/kg and rocuronium 0.3–0.6 mg/kg.

### Maintenance

sevoflurane and fentanyl, with a total fentanyl dose of 3–5 µg/kg.

All patients received a nondepolarizing neuromuscular blocking agent (rocuronium). Depth of anesthesia was not monitored using the bispectral index (BIS).

Groups were defined by whether galantamine had been administered prophylactically immediately after completion of surgery. During the study period, the groups reflected differences in the clinical practice of two anesthesiologists: one routinely used galantamine to reverse neuromuscular blockade in all patients, whereas the other used it as needed. Younger children predominated in the prophylactic galantamine group; the age distribution is shown in Table 1.

**Table 1.**
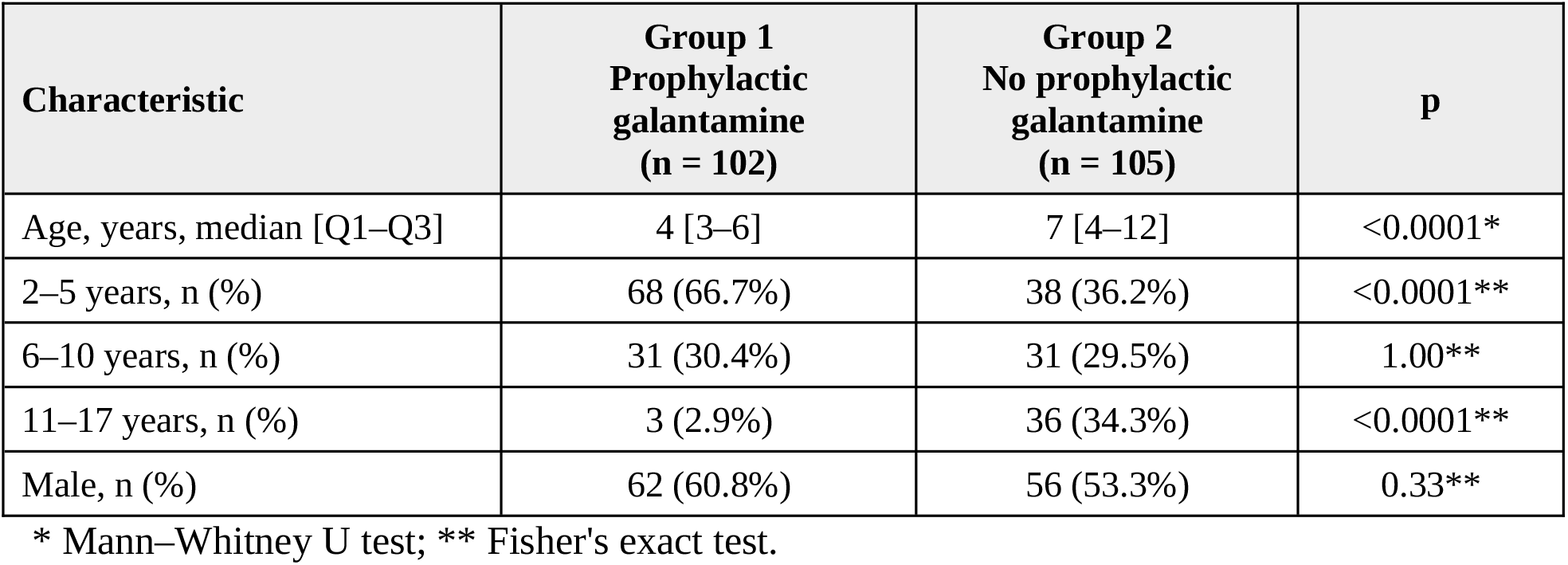
Group characteristics.

| **Characteristic** | **Group 1<br>Prophylactic galantamine<br>(n = 102)** | **Group 2<br>No prophylactic galantamine<br>(n = 105)** | **p** |
|---|---|---|---|
| Age, years, median [Q1–Q3] | 4 [3–6] | 7 [4–12] | <0.0001* |
| 2–5 years, n (%) | 68 (66.7%) | 38 (36.2%) | <0.0001** |
| 6–10 years, n (%) | 31 (30.4%) | 31 (29.5%) | 1.00** |
| 11–17 years, n (%) | 3 (2.9%) | 36 (34.3%) | <0.0001** |
| Male, n (%) | 62 (60.8%) | 56 (53.3%) | 0.33** |

\* Mann–Whitney U test; \*\* Fisher's exact test.

**Table 2.** PAED scores exceeding the primary and exploratory thresholds (n = 207)

| **Outcome** | **Group 1<br>Prophylactic galantamine<br>(n = 102)** | **Group 2<br>No prophylactic galantamine<br>(n = 105)** | **p** |
|---|---|---|---|
| PAED > 10 | 0 (0%) | 19 (18.1%) | <0.0001 |
| PAED > 12<br>Exploratory analysis | 0 (0%) | 14 (13.3%) | <0.0001 |

Group 1 — prophylactic galantamine (n = 102): galantamine was administered intravenously immediately after completion of surgery to reverse neuromuscular blockade in accordance with its approved indication in Russia (“as an antagonist of nondepolarizing muscle relaxants”; current marketing authorization Лп-№ (012692)-(PГ-RU)) [23]. Doses recorded in the final worksheet ranged from 1 to 10 mg, with a median of 2.88 [2.50–4.10] mg. A dose of 2.5 mg was recorded in 28 cases and 5 mg in 11; other doses varied individually.

Group 2 — no prophylactic galantamine, with administration as needed (n = 105): routine reversal of neuromuscular blockade was not performed. Galantamine was administered therapeutically after the PAED score had been recorded when residual neuromuscular blockade, delayed emergence or delirium was suspected. Its use to treat established emergence delirium was outside the approved indications [23].

Dosing was based on our prior clinical experience with Nivalin; we used 0.25–0.3 mg/kg as a dose calculation guide. The Nivalin prescribing information specifies an intravenous dose range of 0.03–0.08 mg/kg (0.25–1 mg) for children aged 1–2 years and 0.03–0.28 mg/kg for children older than 3 years. Doses are selected according to body weight, symptom severity and individual tolerability [24]. Our dose calculation guide describes our clinical practice and does not represent a single labeled regimen applicable to all ages. Actual administered doses in this series were recorded in milligrams.

Incomplete recovery of muscle activity was noted in some episodes of impaired emergence, raising suspicion of residual neuromuscular blockade. In these cases, galantamine was administered to reverse the suspected blockade and improve interaction and behavior. Residual blockade was assessed clinically, without quantitative neuromuscular monitoring.

During recovery from anesthesia, children were observed in the operating room until orientation had returned and they were reunited with their parents. During subsequent ward observation, meaningful interaction between the child and parent was the main clinical indicator. An Aldrete score ≥9 was the criterion for readiness for transfer. All patients received intraoperative paracetamol 15 mg/kg for analgesia. Ondansetron and dexamethasone were combined for prophylaxis of postoperative nausea and vomiting [25], at doses of 0.1 mg/kg and 0.2 mg/kg, respectively. Ibuprofen was used for pain on the ward. When emergence was impaired, the attending anesthesiologist decided whether to administer galantamine. Sedatives were not used to treat delirium.

Emergence delirium was assessed using the PAED scale [26]. Clinical observations made after extubation in the operating room, before treatment for impaired emergence, were recorded in the medical charts. When the analytical worksheet was prepared, total PAED scores were checked against these descriptions and corrected where discrepancies were identified. The primary outcome was PAED > 10 in the final worksheet. The clinical presentation was considered when interpreting this outcome: episodes involving motor agitation and impaired interaction were described separately from failure to awaken.

During retrospective data extraction, vomiting and retching were considered together. Either event during the first postoperative hour was coded as “yes” in the worksheet column labeled “vomiting”.

Analyses of the full sample of 207 patients were performed in Python 3. Age was analyzed in completed years. Quantitative data are presented as median [Q1–Q3] and were compared using a two-sided Mann–Whitney U test with asymptotic calculation, tie correction and continuity correction. Categorical data were compared using a two-sided Fisher’s exact test. An exact 95% confidence interval (CI) for the frequency of PAED > 10 in the prophylactic group was calculated using the Clopper–Pearson method. The 95% CI for the unadjusted between-group difference in proportions was calculated using Newcombe’s method with Wilson intervals without continuity correction. Additional post hoc analyses assessed the frequency of scores exceeding an alternative threshold of PAED > 12 [27] and the primary threshold within the age subgroups 2–5, 6–10 and 11–17 years. These additional comparisons were exploratory; no adjustment for multiple comparisons was applied. Recorded emetic events and the frequency of galantamine administration as needed were also analyzed. Differences were considered statistically significant at p < 0.05.

## RESULTS

Patient characteristics are presented in Table 1.

In the group receiving routine reversal of neuromuscular blockade with galantamine, none of the 102 children had PAED > 10 (0%; exact 95% CI, 0–3.6%). The median PAED score was 7 [6–8], with a range of 4–9. Isolated episodes of brief crying were noted.

In the group without routine reversal, PAED > 10 was recorded in 19 of 105 children (18.1%). The median PAED score was 8 [6–9]; the between-group difference in PAED score distributions was statistically significant (p = 0.0015, Mann–Whitney U test). Scores in children meeting the outcome criterion ranged from 11 to 18. The unadjusted difference in proportions (group 1 minus group 2) was −18.1 percentage points (95% CI, −26.5 to −10.9). At the stricter threshold of PAED > 12 [27], the respective frequencies were 0 of 102 and 14 of 105 (13.3%, p < 0.0001).

The groups differed in age distribution: median age was 4 [3–6] years in the prophylactic galantamine group and 7 [4–12] years in the group without prophylactic administration (p < 0.0001, Mann–Whitney U test). Children aged 2–5 years accounted for 66.7% and 36.2% of the respective groups. No statistically significant difference in sex distribution was found (p = 0.33, Fisher’s exact test). Among children aged 2–5 years, PAED > 10 was recorded in 9 of 38 without prophylactic administration (23.7%) versus 0 of 68 with prophylactic administration (p < 0.0001). The corresponding figures were 4 of 31 (12.9%) versus 0 of 31 in the 6–10-year subgroup (p = 0.11), and 6 of 36 (16.7%) versus 0 of 3 in the 11–17-year subgroup (p = 1.00). Thus, the association persisted in children aged 2–5 years; no statistically significant differences were found in the other age subgroups.

Among the 19 children with PAED > 10, 17 had documented episodes of motor agitation with impaired interaction. In 16 records, lack of interaction was explicitly noted; in the remaining child, disorientation, failure to fix the gaze, screaming and motor agitation were described. In all 17 cases, intravenous galantamine was administered in the operating room at doses of 2.5–10 mg, with a median of 5.8 [4.0–7.0] mg; resolution of the episode was documented in every case. Motor agitation decreased and meaningful interaction returned shortly after administration. Based on our clinical observations, interaction returned more rapidly after galantamine administration during established delirium than after prophylactic administration.

One other child with PAED > 10 had not regained consciousness, interaction or purposeful activity; motor agitation was absent. The condition was considered a suspected comatose form of CAS with possible residual neuromuscular blockade. Galantamine 10 mg was administered to restore consciousness and reverse the suspected blockade; the worksheet recorded PAED 11. Another child, who did not receive galantamine, had PAED 12 and “no” in the clinical symptoms column. Both cases were included in the count of PAED scores > 10 but were not classified among the 17 episodes of delirium with agitation.

Two further children received galantamine for delayed emergence with persisting unconsciousness; the worksheet recorded PAED scores of 10 and 0 and doses of 7 and 20 mg, respectively. Overall, 20 of 105 children received galantamine as needed.

With ondansetron and dexamethasone prophylaxis, emetic events during the first postoperative hour were recorded in 18 of 102 children (17.6%) in the prophylactic galantamine group and 23 of 105 (21.9%) in the group without prophylactic galantamine (p = 0.49). No serious adverse events were documented.

## DISCUSSION

This retrospective observational study tested a clinical hypothesis developed from observations made in 2023–2025. The findings provide a basis for further investigation of galantamine as a potential agent for preventing emergence delirium in children.

In our cohort, no PAED scores > 10 were recorded following prophylactic reversal of neuromuscular blockade with galantamine (0 of 102; 95% CI, 0–3.6%), whereas this threshold was exceeded in 19 of 105 children (18.1%) without prophylactic administration. This association is consistent with the cholinergic hypothesis [22].

Clinically, we distinguished agitation with loss of orientation and interaction from crying directed toward another person, with preserved responses to requests and comforting. The importance of this distinction has been discussed in studies of PAED, pain and behavior during emergence [28, 29, 30]. Delirium can occur despite effective analgesia [31].

The restoration of interaction after galantamine is consistent with the observations of M. S. Danilov and colleagues: in the comatose form of CAS, awakening usually occurred within 30–90 seconds after administration, whereas achievement of a train-of-four (TOF) ratio of 90% during neuromuscular block reversal took a mean of 13.8 ± 8.2 minutes [10]. The possibility of restoring interaction without additional sedation is of interest.

Two approaches to galantamine timing were used in clinical practice: administration immediately after completion of surgery, as used from the outset in the prophylactic group, and administration at the first signs of motor activity. In subsequent clinical practice after the study period, preference was given to administration immediately after completion of surgery: emergence could occur very rapidly when galantamine was administered at the first signs of motor activity, whereas with earlier administration it appeared more gradual. These subsequent observations were not included in the study dataset; the two timings were not formally compared, and the optimal timing remains to be established.

When comparing studies, differences in the outcomes and assessment methods should be considered. PAED separately scores five items: eye contact, purposeful actions, awareness of surroundings, restlessness and inconsolability [26]. This provides a more detailed characterization of the child’s condition, encompassing interaction and awareness of surroundings alongside behavioral agitation. In the Cole and Watcha scales, behavior is assigned a single ordinal category [32, 33, 35].

Funk and colleagues found no significant advantage of physostigmine over placebo in treating agitation in preschool children; the Cole scale used in that study differed from PAED [32, 33]. Cozanitis and colleagues found no advantage of galantamine over neostigmine in memory outcomes in older patients and reported more nausea, vomiting and dysphoria [34].

In the proceedings of the 19th World Congress of Anaesthesiologists (WCA 2026; 15–19 April 2026; Marrakech, Morocco), Pediashov and Gritsan reported a study of prophylactic galantamine involving 80 children undergoing adenoidectomy or adenotonsillectomy [35]. Emergence agitation was assessed using the Watcha scale. Their study and the present study were conducted at the same hospital using separate patient cohorts. Our study used PAED and additionally described treatment of established delirium with galantamine, with documented restoration of meaningful interaction without sedatives.

### Study limitations

The study is based on retrospective data from two anesthesiologists in one department. The groups differed in age (medians of 4 and 7 years) and clinical management, limiting causal interpretation. Assessments were performed by the treating anesthesiologist without blinding. The analytical worksheet contained total PAED scores without separate scores for the five items.

Neuromuscular recovery was assessed clinically. Time to awakening and restoration of interaction were not quantified. The potential contribution of galantamine’s central effects requires separate investigation.

Vomiting and retching were analyzed together during the first hour in the presence of antiemetic prophylaxis. Safety assessment was limited by the completeness of the records.

### Prospects for studying galantamine as an agent promoting emergence

The observations of restored interaction suggest that galantamine could be considered a candidate agent for promoting emergence from anesthesia. Testing this hypothesis requires measurement of time to eye opening, following requests, restoration of orientation and spontaneous breathing, together with neuromuscular and safety monitoring.

## CONCLUSION

### 1. Prophylactic use

With routine reversal of neuromuscular blockade using galantamine, PAED > 10 was not recorded in any of the 102 children. In the group without prophylactic administration, this threshold was exceeded in 19 of 105 children (18.1%).

### 2. Treatment of established delirium

Galantamine was administered in 17 episodes of delirium with motor agitation and impaired interaction. In all 17 cases, resolution of the episode with restoration of meaningful interaction was documented after administration. Sedatives were not used to treat delirium.

These findings from a retrospective observational study support further investigation of both prophylactic and therapeutic galantamine use for emergence delirium in children.

## ADDITIONAL INFORMATION

### Author contributions

Andrey A. Neverov: study concept and design, co-development of the clinical method of galantamine use during emergence from anesthesia, data collection and processing (control group), statistical analysis, and manuscript writing.

Ekaterina A. Kairysh: co-development of the clinical method of galantamine use during emergence from anesthesia, data collection and processing (prophylactic galantamine group), and manuscript editing. Both authors approved the final version of the manuscript.

### Data availability

Anonymized study data are available from the authors upon reasonable request.

### Ethical approval

The study was approved by the Local Ethics Committee of Krasnoyarsk Interdistrict Children’s Hospital No. 4 (protocol No. 1, 24 February 2026) as a retrospective analysis of anonymized routine clinical data. The study involved no additional interventions, tests or changes in treatment. Informed consent from parents or legal representatives for anesthesia was obtained before surgery in accordance with standard procedures. The Local Ethics Committee waived the requirement for separate informed consent from parents or legal representatives for the use of children’s medical data in this retrospective study.

### Use of artificial intelligence

Artificial intelligence tools assisted with statistical calculations, translation and manuscript preparation. The authors are responsible for checking the calculations, sources and final text, and for interpreting the results.

## REFERENCES

1. Vlajkovic GP, Sindjelic RP. Emergence delirium in children: many questions, few answers. Anesth Analg. 2007;104(1):84–91. 10.1213/01.ane.0000250914.91881.a8

2. Aldecoa C, Bettelli G, Bilotta F, et al. European Society of Anaesthesiology evidence-based and consensus-based guideline on postoperative delirium. Eur J Anaesthesiol. 2017;34(4):192–214. 10.1097/EJA.0000000000000594

3. Liu SL, Bai XD, Xu T, et al. Prevalence and influencing factors of postoperative delirium in children undergoing low-risk surgeries: a meta-analysis. J Pediatr Nurs. 2026;89:38–49. 10.1016/j.pedn.2026.04.005

4. Chen YC, Foster J, Wang ML, et al. Global prevalence and risk factors of emergence delirium in pediatric patients undergoing general anesthesia: a systemic review and meta-analysis. J Pediatr Nurs. 2024;77:74–80. 10.1016/j.pedn.2024.03.010

5. Likhvantsev VV, Ulitkina ON, Rezepov NA. [Postoperative delirium: what is new in the ESA-2017 guideline?]. Vestnik Anesteziologii i Reanimatologii. 2017;14(2):41–47. Russian. 10.21292/2078-5658-2017-14-2-41-47

6. Kolodyazhnaya VI, Aleksandrovich YuS, Grechanyi SV. [Emergence excitation, agitation and delirium following sevoflurane anesthesia in children]. Anesteziologiya i Reanimatologiya. 2023;(5):74–80. Russian. 10.17116/anaesthesiology202305174

7. Wong DDL, Bailey CR. Emergence delirium in children. Anaesthesia. 2015;70(4):383–387. 10.1111/anae.13043

8. Dahmani S, Delivet H, Hilly J. Emergence delirium in children: an update. Curr Opin Anaesthesiol. 2014;27(3):309–315. 10.1097/ACO.0000000000000076

9. Mason KP. Paediatric emergence delirium: a comprehensive review and interpretation of the literature. Br J Anaesth. 2017;118(3):335–343. 10.1093/bja/aew477

10. Danilov MS, Lebedinskii KM, Kurapeev IS. Central anticholinergic syndrome following general anaesthesia: prevention and treatment with galantamine. Herald of North-Western State Medical University named after I.I. Mechnikov. 2018;10(1):43–48. Russian. 10.17816/mechnikov201810143-48

11. Heinrich M, Teoh HL. Galanthamine from snowdrop—the development of a modern drug against Alzheimer’s disease from local Caucasian knowledge. J Ethnopharmacol. 2004;92(2–3):147–162. 10.1016/j.jep.2004.02.012

12. Derezhian A, Krusteva E. [Our experiences with decurarizing effects of nivaline]. Khirurgiia (Sofiia). 1960;13:272–274. Bulgarian. https://pubmed.ncbi.nlm.nih.gov/13816031/

13. Stoyanov E, Vulchanova S. The clinical application of Nivalin as an antidote of curare. Nauchni Tr Vissh Med Inst Sofiia. 1963;42:45–48. https://pubmed.ncbi.nlm.nih.gov/14128111/

14. Wislicki L. Nivalin (galanthamine hydrobromide), an additional decurarizing agent. Some introductory observations. Br J Anaesth. 1967;39(12):963–968. 10.1093/bja/39.12.963

15. Chakalova E, Marinova M, Srebreva M, Anastasov D, Ploskov K. [Attempt to eliminate residual somnolence and disorientation with nivaline after anesthesia with ketalar and diazepam for minor obstetrical and gynecologic surgery]. Akush Ginekol (Sofiia). 1987;26(3):28–31. Bulgarian. https://pubmed.ncbi.nlm.nih.gov/3631427/

16. Volkov VP. [Anticholinergic syndrome: a literature review]. Psikhicheskie Rasstroystva v Obshchey Meditsine. 2012;(3):52–58. Russian. https://con-med.ru/magazines/psikhicheskie_rasstroystva_v_obshchey_meditsine/psikhicheskie_rasstroystva_v_obshchey_meditsine-03-2012/antikholinergicheskiy_sindrom_obzor_literatury_/

17. Schneck HJ, Rupreht J. Central anticholinergic syndrome (CAS) in anesthesia and intensive care. Acta Anaesthesiol Belg. 1989;40(3):219–228. https://pubmed.ncbi.nlm.nih.gov/2683549/

18. Link J, Papadopoulos G, Dopjans D, Guggenmoos-Holzmann I, Eyrich K. Distinct central anticholinergic syndrome following general anaesthesia. Eur J Anaesthesiol. 1997;14(1):15–23. 10.1046/j.1365-2346.1997.00004.x

19. Cook B, Spence AA. Post-operative central anticholinergic syndrome. Eur J Anaesthesiol. 1997;14(1):1–2. 10.1046/j.1365-2346.1997.00074.x

20. Danilov MS, Lebedinskii KM. [Central anticholinergic… syndrome?]. Anesteziol Reanimatol. 2015;60(6):75–78. Russian. https://pubmed.ncbi.nlm.nih.gov/27025142/

21. Baraka A, Harik S. Reversal of central anticholinergic syndrome by galanthamine. JAMA. 1977;238(21):2293–2294. 10.1001/jama.1977.03280220061024

22. Hshieh TT, Fong TG, Marcantonio ER, Inouye SK. Cholinergic deficiency hypothesis in delirium: a synthesis of current evidence. J Gerontol A Biol Sci Med Sci. 2008;63(7):764–772. 10.1093/gerona/63.7.764

23. VIFITECH. Galantamine, solution for intravenous and subcutaneous administration, 1 mg/mL and 5 mg/mL: prescribing information. Marketing authorization Лп-№(012692)-(PГ-RU), dated 3 December 2025. Reproduced prescribing information: https://medvestnik.ru/ls/a16ca2dd-9512-495c-8c8f-e577d246e5b4.html (accessed 6 November 2026). Russian.

24. Sopharma. Nivalin, solution for injection: prescribing information. Marketing sauthorization п N008777. https://medi.ru/instrukciya/nivalin_5387/ (accessed 5 November 2026). Russian.

25. Gan TJ, Belani KG, Bergese S, et al. Fourth Consensus Guidelines for the Management of Postoperative Nausea and Vomiting. Anesth Analg. 2020;131(2):411–448. 10.1213/ANE.0000000000004833

26. Sikich N, Lerman J. Development and psychometric evaluation of the pediatric anesthesia emergence delirium scale. Anesthesiology. 2004;100(5):1138–1145. 10.1097/00000542-200405000-00015

27. Klabusayová E, Musilová T, Fabián D, et al. Incidence of emergence delirium in the pediatric PACU: prospective observational trial. Children (Basel). 2022;9(10):1591. 10.3390/children9101591

28. Somaini M, Engelhardt T, Fumagalli R, Ingelmo PM. Emergence delirium or pain after anaesthesia—how to distinguish between the two in young children: a retrospective analysis of observational studies. Br J Anaesth. 2016;116(3):377–383. 10.1093/bja/aev552

29. Ringblom J, Wåhlin I, Proczkowska M, Korhonen L, Årestedt K. Measurement Properties of the Pediatric Anesthesia Emergence Delirium Scale: A Confirmatory Factor Analysis-Based Study. Paediatr Anaesth. 2025;35(2):155–162. 10.1111/pan.15046

30. O’Brien JM, McKay WPS, McDonald M. Identifying behaviors that characterize emergence delirium: An observational study. Paediatr Anaesth. 2023;33(8):631–635. 10.1111/pan.14678

31. Locatelli BG, Ingelmo PM, Emre S, Meroni V, Minardi C, Frawley G, et al. Emergence delirium in children: a comparison of sevoflurane and desflurane anesthesia using the Paediatric Anesthesia Emergence Delirium scale. Paediatr Anaesth. 2013;23(4):301–308. 10.1111/pan.12038

32. Funk W, Hollnberger H, Geroldinger J. Physostigmine and anaesthesia emergence delirium in preschool children: a randomized blinded trial. Eur J Anaesthesiol. 2008;25(1):37–42. 10.1017/S0265021507001159

33. Häusler JM. Physostigmin und postoperatives Delir bei Vorschulkindern: eine randomisierte Doppelblindstudie [Dissertation]. Regensburg: Universität Regensburg; 2013. 10.5283/epub.27184

34. Cozanitis D, Keinonen M, Maunuksela EL. Effect on cognition of galanthamine administered for neuromuscular block reversal in octogenarians undergoing cataract surgery. Anaesthesiol Intensive Ther. 2012;44(2):76–80. https://pubmed.ncbi.nlm.nih.gov/22992965/

35. Pediashov V, Gritsan A. Effectiveness of Galantamine for the Prevention of Emergence Agitation in Children After Adeno-or Adenotonsillectomy [conference abstract]. 19th World Congress of Anaesthesiologists; 15–19 April 2026; Marrakech, Morocco. Abstract PA.E6.01.01. https://pdf-program.abstractserver.com/?congress=wca2026

